# Association of lipid-lowering drugs with COVID-19 outcomes: A Mendelian Randomization study

**DOI:** 10.1101/2021.07.20.21260813

**Authors:** Wuqing Huang, Jun Xiao, Jianguang Ji, Liang-Wan Chen

**Affiliations:** Department of Epidemiology and Health Statistics, School of Public Health, Fujian Medical University, Fuzhou, Fujian, China; Department of Cardiovascular Surgery, Fujian Medical University Union Hospital, Fuzhou, China; Center for Primary Health Care Research, Department of Clinical Sciences Malmö, Lund University, Sweden

**Keywords:** lipid-lowering drugs, COVID-19, Mendelian randomization

## Abstract

**Background:** Lipid metabolism plays an important role in viral infections. Large cohort study suggested a protective potential of lipid-lowering drugs in COVID-19 outcomes, but the nature of observational study precludes it to draw a causal inference.

**Objectives:** To assess the causal effect of lipid-lowering drugs (HMGCR inhibitors, PCSK9 inhibitors and NPC1L1 inhibitors) on COVID-19 outcomes using 2-sample Mendelian Randomization (MR) study.

**Methods:** We used two kinds of genetic instruments to proxy the exposure of lipid-lowering drugs, including expression quantitative trait loci (eQTLs) of drugs target genes, and genetic variants within or nearby drugs target genes associated with low-density lipoprotein (LDL) cholesterol from genome-wide association study (GWAS). GWASs of COVID-19 outcomes (susceptibility, hospitalization and very severe disease) were obtained from the COVID-19 Host Genetics Initiative. Summary-data-based MR (SMR) and inverse-variance weighted MR (IVW-MR) were used to calculate the effect estimates.

**Results:** SMR analysis found that a higher expression of HMGCR was associated with a higher risk of COVID-19 hospitalization (OR=1.38, 95%CI=1.06-1.81; P=0.019). Similarly, IVW-MR analysis observed a positive association between HMGCR-mediated LDL cholesterol and COVID-19 hospitalization (OR=1.32, 95%CI=1.00-1.74; P=0.049). No consistent evidence from both SMR and IVW-MR analyses was found for the association of HMGCR inhibitors with COVID-19 susceptibility or very severe disease, or for the association of PCSK9 inhibitors and NPC1L1 inhibitor with COVID-19 outcomes.

**Conclusions:** In this 2-sample MR study, we found potential causal evidence that HMGCR inhibitors could reduce the risk of COVID-19 hospitalization. Further research is needed to explore the therapeutic role of statins for COVID-19.

## Introduction

The COVID-19 pandemic has caused millions of infections and deaths, which is caused by a novel coronavirus, severe acute respiratory syndrome coronavirus 2 (SARS-CoV-2). Lacking of drugs specifically targeted to SARS-CoV-2 infection has led to a great interest to identify drugs that can be repurposed to reduce the infection and mortality of the disease. Available studies have suggested an important role of lipid metabolism in viral infections, including in the pathogenesis of SARS-CoV-2 infection (1). The plausible mechanisms include the involvement of host lipids in the virus life cycle, the influence of cholesterol on the immune cell functions, interfering with the mevalonate pathway, and so on (1). Such evidence indicates the potential protective effect of lipid-lowering drugs against COVID-19. HMG-CoA Reductase (HMGCR) inhibitors, known as statins, are the most commonly-used class of lipid-lowering drugs, which have a couple of predominant merits, such as the well-proven safety, low cost, and pleiotropic effects. Therefore, a number of observational studies have investigated the association between statins and COVID-19 outcomes, but generating mixed results (2-6). What’s more, confounding bias and reverse causation cannot be avoided in most of these studies.

Mendelian randomization (MR) study uses genetic variants as an instrument to perform causal inference between an exposure and an outcome, which could determine whether an observational association is consistent with a causal effect (7). Confounding bias can be minimized in MR study because genetic variants are randomly assigned to the individual at birth. Similarly, reverse causation can be avoided because genetic variants are assigned prior to the development of disease.

Therefore, we performed 2-sample MR analysis in this study to test the association of several FDA-approved lipid-lowering drugs (HMGCR inhibitors, PCSK9 inhibitors and NPC1L1 inhibitors) with COVID-19 outcomes (susceptibility, hospitalization and very severe disease).

## Methods

### Study design

This 2-sample MR study is based on publicly available summary-level data from genome-wide association studies (GWASs) and expression quantitative trait loci (eQTLs) studies (**Supplementary Table 1**). All these studies had been approved by the relevant institutional review boards and participants had provided informed consents.

**Table 1.** Information of genetic instruments.

| Exposure | Genetic instruments |  |
| --- | --- | --- |
|  | Genetic variants associated with mRNA expression levels (eQTLs) | Genetic variants associated with LDL cholesterol level |
| HMGCR inhibitors | 168 common cis-eQTLs (MAF>1%) in blood for HMGCR gene ($P < 5.0 \times 10^{-8}$), top SNP: rs6453133 | 7 common SNPs (MAF>1%) in low linkage disequilibrium ($r^2 < 0.30$), associated with LDL cholesterol ($P < 5.0 \times 10^{-8}$), located within $\pm 100$ kb windows from HMGCR region |
| PCSK9 inhibitors | 24 common cis-eQTLs (MAF>1%) in blood for PCSK9 gene ($P < 5.0 \times 10^{-8}$), top SNP: rs472495 | 12 common SNPs (MAF>1%) in low linkage disequilibrium ($r^2 < 0.30$), associated with LDL cholesterol ($P < 5.0 \times 10^{-8}$), located within $\pm 100$ kb windows from PCSK9 region |
| NPC1L1 inhibitors | 11 common cis-eQTLs (MAF>1%) in adipose subcutaneous tissue for NPC1L1 gene ($P < 5.0 \times 10^{-8}$), top SNP: rs41279633 | 3 common SNPs (MAF>1%) in low linkage disequilibrium ($r^2 < 0.30$), associated with LDL cholesterol ($P < 5.0 \times 10^{-8}$), located within $\pm 100$ kb windows from NPC1L1 region |
| **Statistical analyses** |  |  |
| Primary analysis | Summary-data-based Mendelian Randomization | Inverse-variance weighted Mendelian Randomization |
| Sensitivity analyses | F-statistic<br>Positive control analysis (LDL cholesterol used as outcome)<br>Linkage disequilibrium test: HEIDI test<br>Horizontal pleiotropy test: SMR association between expression of adjacent genes and outcome | F-statistic<br>Positive control analysis (coronary heart disease used as outcome)<br>Heterogeneity test: Cochran Q test<br>Horizontal pleiotropy test: MR-Egger regression, MR-PRESSO test |

Abbreviations and Acronyms: eQTLs, expression quantitative trait loci; GWAS, genome-wide association study; HEIDI, heterogeneity in dependent instruments; HMGCR, HMG-CoA reductase; IVW-MR, inverse-variance weighted Mendelian Randomization; LDL, low-density lipoprotein; MR-PRESSO, Mendelian Randomization RESidual Sum and Outlier; MAF, minor allele frequency; NPC1L1, Niemann-Pick C1-Like 1; PCSK9, Proprotein convertase subtilisin/kexin type 9; SNP, single-nucleotide polymorphism; SMR, summary-data-based Mendelian Randomization.

### Selection of genetic instruments

Three classes of FDA-approved lipid-lowering drugs were included as exposures in this study: HMGCR inhibitors (i.e., statins), proprotein convertase subtilisin/kexin type 9 (PCSK9) inhibitors (i.e., alirocumab and evolocumab), and niemann-Pick C1 like 1 (NPC1L1) inhibitor (i.e., ezetimibe).

As shown in **Table 1**, we used available eQTLs for drugs target genes (i.e., HMGCR, PCSK9, NPC1L1) as the proxy of exposure to each lipid-lowering drug. The eQTLs summary-level data was obtained from eQTLGen Consortium (https://www.eqtlgen.org/) or GTEx Consortium V8 (https://gtexportal.org/home/). We identified common (minor allele frequency (MAF) >1%) eQTLs single-nucleotide polymorphisms (SNPs) significantly (p<5.0 × 10^−8^) associated with the expression of HMGCR or PCSK9 in blood, and the expression of NPC1L1 in adipose subcutaneous tissue as there are no eQTLs in blood or other tissues available at a significance level for NPC1L1. Only cis-eQTLs were included to generate genetic instruments in this study, which were defined as eQTLs within 1 Mb on either side of the encoded gene.

Secondly, to validate the observed association using the eQTLs as an instrument, we additionally proposed an instrument by selecting SNPs within 100 kb windows from target gene of each drug that were associated with LDL cholesterol level at a genome-wide significance level (p<5.0 × 10^−8^) to proxy the exposure of lipid-lowering drugs. A GWAS summary data of LDL cholesterol levels from the Global Lipids Genetics Consortium was used to identify these SNPs, where only common SNPs (MAF>1%) were included (8). Seven SNPs within 100 kb windows from HMGCR gene were selected for proxying HMGCR inhibitors, 12 SNPs from PCSK9 gene identified for PCSK9 inhibitors, and three SNPs from NPC1L1 gene selected for NPC1L1 inhibitor. To maximize the strength of the instrument for each drug, SNPs used as instruments were allowed to be in low weak linkage disequilibrium (r^2^ <0.30) with each other.

### Outcome sources

GWAS summary-level data for COVID-19 outcomes were obtained from the COVID-19 Host Genetics Initiative V4 (https://www.covid19hg.org/) (9). The study population was restricted to individuals with European ancestry, including meta-analyses of GWASs containing up to 22 cohorts from 11 countries. GWAS from these cohorts used a model adjusted for age, sex, age × age, age × sex, genetic principal components, and study-specific covariates. A COVID-19 case was confirmed by lab or self-reported infections, or electronic health records of infections. The susceptibility outcome was measured by comparing COVID-19 cases and controls who did not have a history of COVID-19. The hospitalized outcome was measured by comparing COVID-19 hospitalized cases and controls who were never admitted to the hospital due to COVID-19, including individuals without COVID-19. The severe disease outcome was measured by comparing COVID-19 cases who died or required respiratory support and controls without severe COVID-19, including individuals without COVID-19. We included individuals without COVID-19 as controls for all outcomes to decrease collider bias and allow for population-level comparisons (10,11).

### Statistical analyses

#### Primary MR analysis

Summary-data-based MR (SMR) method was applied to generate effect estimates when using eQTLs as an instrument, which investigates the association between the expression level of a gene and outcome of interest using summary-level data from GWAS and eQTL studies (12). Allele harmonization and analysis were performed using SMR software, version 1.03 (https://cnsgenomics.com/software/smr/#Overview). Inverse-variance weighted MR (IVW-MR) method was used to combine effect estimates when using genetic variants associated with LDL cholesterol level as an instrument. Allele harmonization and analysis were conducted using the TwoSampleMR package in R software, version 4.1.0.

#### Sensitivity analysis

The strength of SNPs used as the instrument was assessed using the F-statistic, and we included SNPs with an F-statistic of >10 to minimize weak instrument bias (13). Positive control analyses were performed for validation of both genetic instruments. Since lowering the level of LDL cholesterol is the well-proven effect of lipid-lowering drugs, we thus examined the association of exposures of interest with LDL cholesterol level as positive control study for the instrument from eQTLs. For the instrument from LDL cholesterol GWAS, we performed positive control study by examining the association of exposures of interest with coronary heart disease because coronary heart disease is the main indication of lipid-lowering drugs.

For SMR method, the heterogeneity in dependent instruments (HEIDI) test was used to test if the observed association between gene expression and outcome was due to a linkage scenario, which was performed in the SMR software (12). As recommended by the authors, the HEIDI test of P<0.01 indicates that association is probably due to linkage (12). One SNP could be related to the expression of more than one gene, leading to the presence of horizontal pleiotropy. To assess the risk of horizontal pleiotropy, we identified other nearby genes (within a 1Mb window) with significant association with the genetic instrumental variant, and performed SMR analysis to examine if the expression of these genes was related to the COVID-19 outcomes.

For IVW-MR method, we tested the heterogeneity by using a Cochran Q test, where P<0.05 indicates the evidence of heterogeneity (14). MR-Egger regression and Mendelian Randomization Pleiotropy RESidual Sum and Outlier (MR-PRESSO) analysis were used to assess the potential horizontal pleiotropy of the SNPs used as instrument variants. In MR Egger regression, the intercept term is a useful indication of directional horizontal pleiotropy, where P<0.05 indicates the evidence of horizontal pleiotropy (15). MR-PRESSO analysis can identify horizontal pleiotropic outliers and provide adjusted estimates, where P<0.05 for Global test indicates the presence of horizontal pleiotropic outliers (16). All these analyses were implemented in R software, version 4.1.0.

To account for multiple testing correction, Bonferroni correction was used to adjust the thresholds of significance level, with a strong evidence of P<0.006 (3 exposures and 3 outcomes) and a suggestive evidence of 0.006≤P<0.05.

## Results

### Genetic instruments selection and COVID-19 outcomes

A total of 168, 24, and 11 cis-eQTLs were identified from eQTLGen or GTEx Consortium for drugs target gene HMGCR, PCSK9, and NPC1L1, respectively, and the most significant cis-eQTL SNP was selected as a genetic instrument for target gene of each drug (**Table 1, Supplementary Table 2**). A total of 7, 12, and 3 SNPs within or nearby gene HMGCR, PCSK9, and NPC1L1 were selected from a GWAS summary data of LDL cholesterol levels in the Global Lipids Genetics Consortium, respectively (**Table 1, Supplementary Table 3**). F-statistics for all instrument variants were over 30, suggesting that weak instrument bias can be minimized in our study (**Supplementary Table 2 and 3**). Positive control study showed significant associations between exposure to each drug and LDL cholesterol when using eQTLs-proposed instruments (**Supplementary Table 5**), as well as between exposure to each drug and coronary heart disease when using LDL cholesterol GWAS-proposed instruments (**Supplementary Table 6**), further ensuring the efficacy of the selected genetic instruments.

From COVID-19 GWASs, a total of 14,134 cases and 1,284,876 controls were used to explore the association with COVID-19 susceptibility, 6,406 cases and 902,088 controls for COVID-19 hospitalization, and 3,886 cases and 622,265 controls for COVID-19 severe disease (**Supplementary Table 1**).

### Primary analysis

In **Figure 1 and Supplementary Table 2**, results from SMR analysis found a suggestive evidence for the association of the increased expression of HMGCR gene in blood (equivalent to a 1 standard deviation increase) with the higher risk of COVID-19 susceptibility (OR=1.30, 95%CI=1.05-1.61; P=0.017) and COVID-19 hospitalization (OR=1.38, 95%CI=1.06-1.81; P=0.019), indicating that HMGCR inhibitors can lower the risk of COVID-19 susceptibility and hospitalization. A suggestive evidence was observed regarding the negative association between PCSK9 expression and risk of COVID-19 susceptibility (OR=0.84, 95%CI=0.73-0.97; P=0.02). No significant association was found between the expression of NPC1L1 and COVID-19 outcomes.

**Figure 1.**
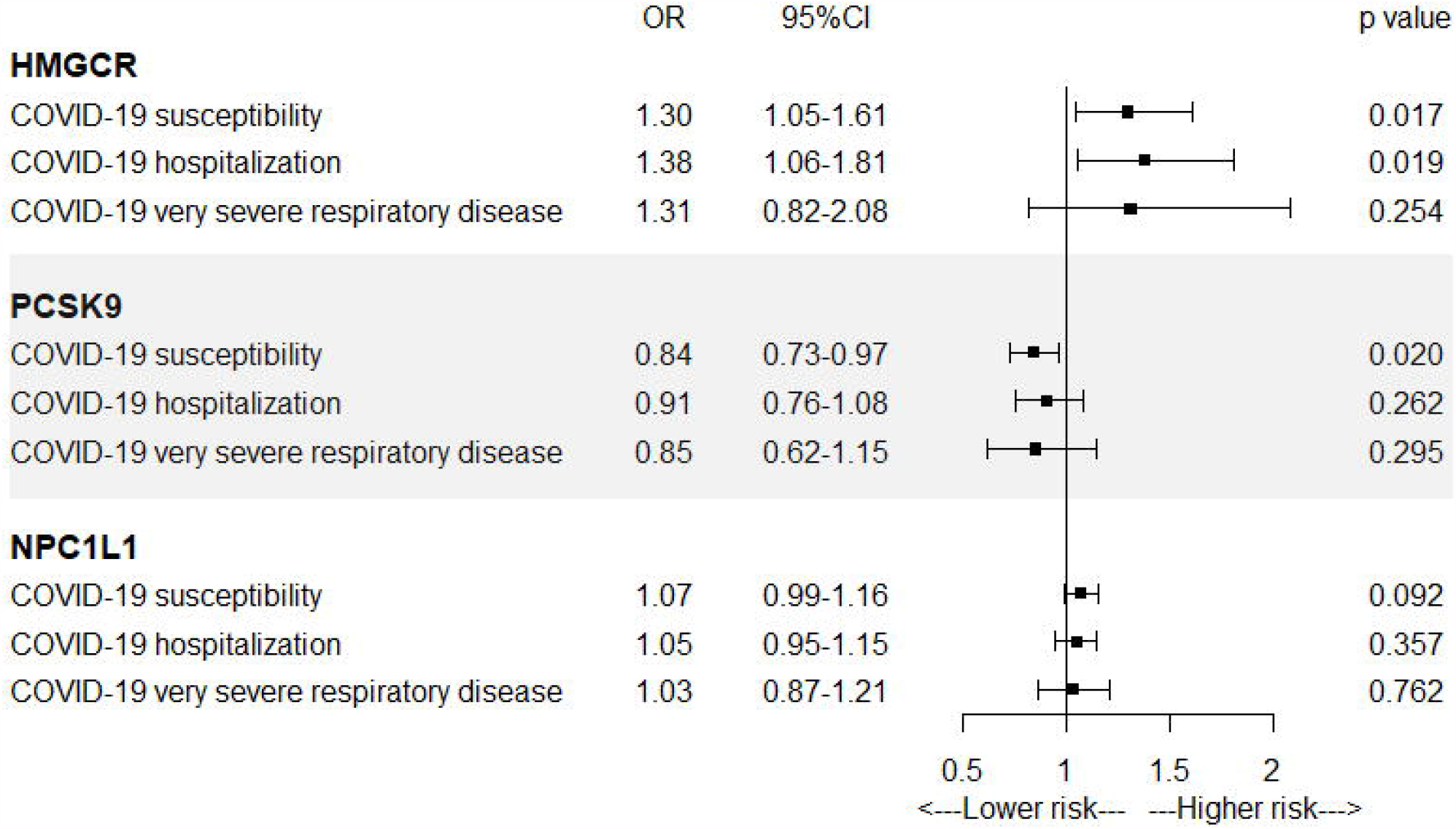
SMR association between expression of gene HMGCR, PCSK9, or NPC1L1 and COVID-19 outcomes. Abbreviations and Acronyms: CI, confidence interval; HMGCR, HMG-CoA reductase; NPC1L1, Niemann-Pick C1-Like 1; OR, odds ratio; PCSK9, Proprotein convertase subtilisin/kexin type 9; SMR, summary-data-based Mendelian Randomization.

In **Figure 2 and Supplementary Table 4**, IVW-MR analysis also found a suggestive evidence for the association between HMGCR-mediated LDL cholesterol (equivalent to a 1-mmol/L increase) and the risk of COVID-19 hospitalization (OR=1.32, 95%CI=1.00-1.74; P=0.049), further supporting the protective effect of HMGCR inhibitors against COVID-19 hospitalization. A strong evidence was observed between NPC1L1-mediated LDL cholesterol and risk of COVID-19 susceptibility (OR=2.02, 95%CI=1.30-3.13; P=0.002). IVW-MR analysis did not provide any evidence for the association between PCSK9-mediated LDL cholesterol and COVID-19 outcomes.

**Figure 2.**
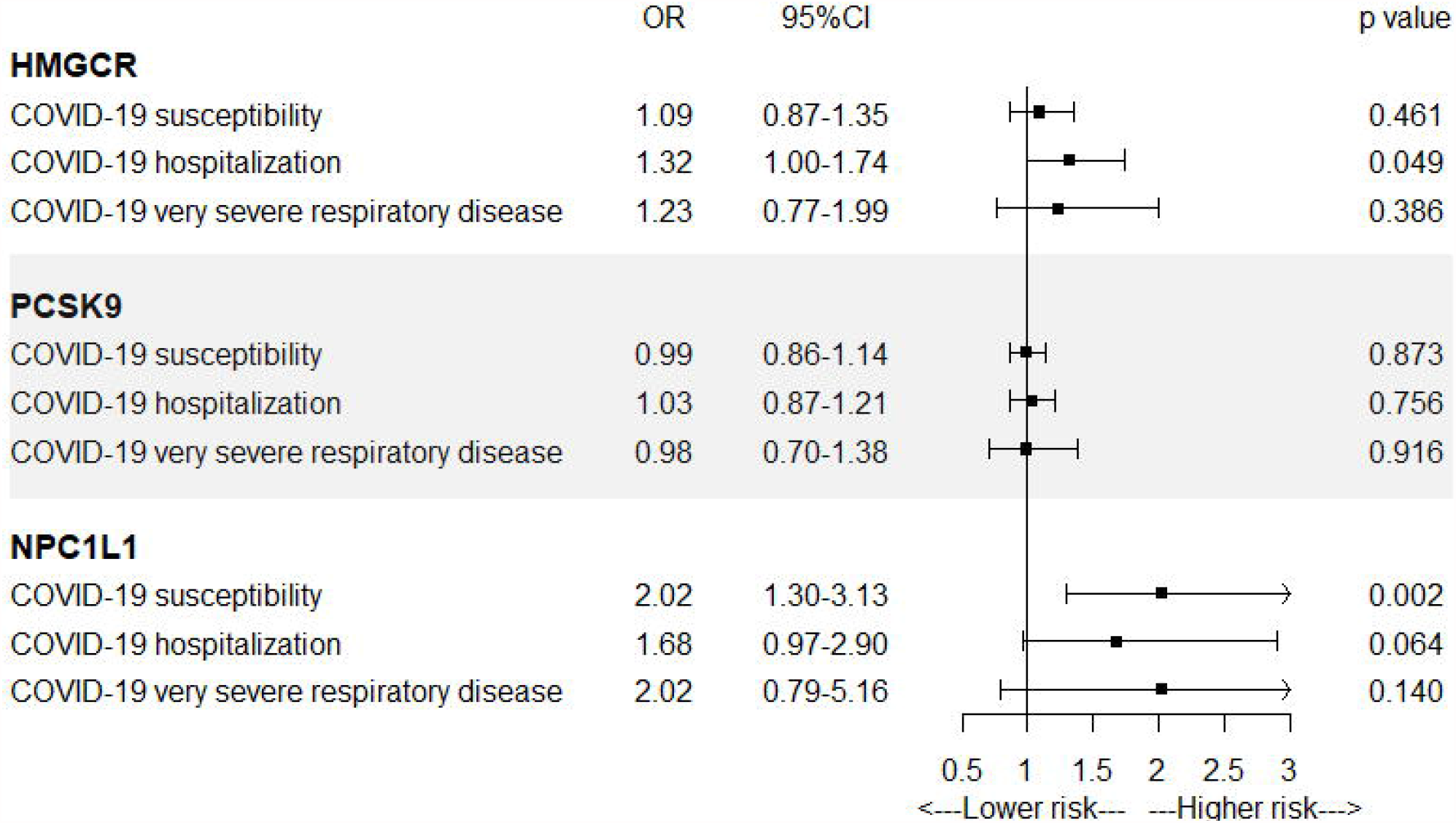
IVW-MR association between LDL cholesterol mediated by gene HMGCR, PCSK9, or NPC1L1 and COVID-19 outcomes. Abbreviations and Acronyms: CI, confidence interval; HMGCR, HMG-CoA reductase; IVW-MR, inverse-variance weighted Mendelian Randomization; NPC1L1, Niemann-Pick C1-Like 1; OR, odds ratio; PCSK9, Proprotein convertase subtilisin/kexin type 9.

### Sensitivity analysis

For SMR analysis, HEIDI test suggested that all observed associations were not due to a linkage (P>0.01), except for the association between HMGCR expression and COVID-19 susceptibility (P=0.009) (**Supplementary Table 2**). We further examine if horizontal pleiotropy was present in the association between HMGCR expression and COVID-19 outcomes by investigating if there was an association between the expression of nearby genes which are significantly associated with the top eQTL SNP (instrument variant) of HMGCR and COVID-19 outcomes. We identified six genes, including HMGCR, the expression of which were associated with the instrument variant (**Supplementary Table 7**). Only four genes have available eQTLs at a genome-wide significance level (p<5.0 × 10^−8^). Among these four genes, only HMGCR expression was significantly related to COVID-19 susceptibility and COVID-19 hospitalization, suggesting a small role of horizontal pleiotropy in the observed associations (**Supplementary Table 8**).

For IVW-MR analysis, Cochran Q test did not found evidence of heterogeneity for all reported results (All P<0.05; **Supplementary Table 4**). Both the intercept term in MR– Egger regression and MR-PRESSO analysis suggested that horizontal pleiotropy played a small role in the reported results (All P<0.05; **Supplementary Table 4**).

## Discussion

The present MR study provided a suggestive evidence regarding the positive association of the HMGCR expression and HMGCR-mediated LDL cholesterol level with the risk of COVID-19 hospitalization, both of which together indicated a protective effect of HMGCR inhibitors against COVID-19 hospitalization (OR _instrument 1_=0.72, 95%CI=0.55-0.95; OR _instrument 2_=0.76, 95%CI=0.57-1.00). We found a suggestive evidence of the negative association between PCSK9 expression and COVID-19 susceptibility, but which was not supported when using LDL cholesterol GWAS as an instrument. A strong evidence was observed for the protective effect of NPC1L1-mediated lower level of LDL cholesterol on COVID-19 susceptibility, but there was no evidence for the association of NPC1L1 expression and COVID-19 outcomes.

Compared to develop a new drug, repurposing an old drug is much more economical and time-saving, in particular, the importance is further highlighted during a pandemic. COVID-19 pandemic has driven a number of studies of drug repurposing (17,18). The role of lipid metabolism in viral infections has raised the interest regarding the possibility of repurposing lipid-lowering drugs as anti-COVID-19 agents (1). As one of the most commonly prescribed drugs, statins have received the greatest attention for their pleiotropic effects, including lowering serum cholesterol, anti-inflammatory and immunomodulatory properties and antithrombotic effect, all of which play a role in viral infections (1,17,19). Emerging observational studies have investigated if statins might benefit patients with COVID-19 (2-6). A largest retrospective cohort study including 13,981 patients admitted to hospital due to COVID-19 suggested a significant reduction in 28-day all-cause mortality by 42% in the group with statins than patients without statins (2). A meta-analysis with 8,990 COVID-19 patients also found a 30% lower risk of fatal or severe disease (3). However, a Danish nationwide cohort study with 4,842 COVID-19 patients and a meta-analysis with 3,449 COVID-19 patients did not found the association between statins use and improved COVID-19 outcomes (5,6). In addition, in these studies, considerable differences in clinical characteristics cannot be avoided between patients with and without statins, and causal inference is not allowed due to the retrospective nature of observational studies.

As a genetic epidemiological method, MR study could overcome the limitations of traditional observational studies. In this MR study, we used genetic variants related to HMGCR expression or HMGCR-mediated LDL cholesterol as instruments to proxy the exposure of statins. Both analyses found a suggestive evidence that statins could reduce the risk of COVID-19 hospitalization, rather than COVID-19 susceptibility and very severe outcome. Although strong evidence is lacking, these results provided a causal evidence supporting the finding from the largest cohort study (2), which calls for additional observational studies in different populations, mechanistic studies, and randomized controlled studies to examine its potential effect against COVID-19. Besides, although no association was found between NPC1L1 expression in adipose subcutaneous and COVID-19 outcomes, there was a strong evidence of the association between NPC1L1-mediated LDL cholesterol and COVID-19 susceptibility. The effect of NPC1L1 inhibitor on COVID-19 susceptibility may be worth further studies as well.

### Study strengths

The main strength of our study is the use of genetic instruments to proxy drug exposure, which could minimize confounding bias and avoid reverse causation. Besides, we used two different kinds of genetic instruments to proxy the studied drug, which contributes to validate the effect estimates from each other. And a number of sensitivity analyses have been performed to test the efficacy of genetic instruments and the assumptions of MR study. In addition, we used large eQTL summary data of drugs target genes and GWAS summary data of LDL cholesterol and COVID-19 outcomes, which could ensure statistical power.

### Study limitations

This study has several limitations. Firstly, there are no available eQTLs in blood for NPC1L1, so we were not allowed to explore the association between NPC1L1 expression in blood and COVID-19 outcomes. Besides, there are no available eQTLs in lung for HMGCR, which might provide more convincing evidence of the association between HMGCR inhibitors and COVID hospitalization. Secondly, the effect of statins probably varies between subgroups, for example, it may be more effective in patients with chronic diseases (e.g., coronary heart disease). However, the use of summary-level data did not allow us to perform subgroup analyses, so further MR study with individual-level data is needed to provide more detailed information. Thirdly, the Bonferroni correction for multiple tests suggests that we cannot rule out the false-positive possibility for the finding of the protective effect of statins on COVID hospitalization. Fourthly, confounding bias and/or horizontal pleiotropy cannot be completely excluded although we have performed various sensitivity analyses to test the assumptions of MR study.

## Conclusions

In conclusion, this MR study provided potential causal evidence for the association between statins exposure and the reduced risk of COVID-19 hospitalization. Further research is called to confirm the association and the underlying mechanisms.

## Data Availability

All databases used in this study are publicly available. Covid-19 outcome GWAS summary statistics were downloaded from the Covid-19 HGI website. The eQTL summary statistics were downloaded from eQTLGen Consortium or GTEx Consortium V8.

https://www.covid19hg.org/

https://www.eqtlgen.org/

https://gtexportal.org/home/

## Author Contributors

WH, JX, JJ, and LWC were responsible for the study concept and design. WH and JX acquired the data. WH and JX did the statistical analysis and drafted the manuscript, and all authors revised it for important intellectual content. WH and JX contributed equally to the study.

## Acknowledgments

We thank the patients and investigators who contributed to the eQTLGen Consortium, GTEx Consortium, COVID-19 Host Genetics Initiative, Global Lipids Genetics Consortium and CARDIoGRAMplusC4D Consortium.

## Abbreviations and Acronyms

CI: confidence interval
eQTLs: expression quantitative trait loci
GWAS: genome-wide association study
HEIDI: heterogeneity in dependent instruments
HMGCR: HMG-CoA reductase
IVW-MR: inverse-variance weighted Mendelian Randomization
LDL: low-density lipoprotein
MR-PRESSO: Mendelian Randomization Pleiotropy RESidual Sum and Outlier
MAF: minor allele frequency
NPC1L1: Niemann-Pick C1-Like 1
OR: odds ratio
PCSK9: Proprotein convertase subtilisin/kexin type 9
SNP: single-nucleotide polymorphism
SMR: summary-data-based Mendelian Randomization.

## Perspectives

### COMPETENCY IN MEDICAL KNOWLEDGE

A suggestive evidence indicates that statins use is causally associated with a reduced risk of COVID-19 hospitalization.

### TRANSLATIONAL OUTLOOK

The protective effect of statins should be further confirmed in randomized controlled trials.

### Central Illustrationss

Lipid-lowering drugs and COVID-19 outcomes.

Lipid metabolism plays an important role in the viral infections, indicating the potential to be repurposed against COVID-19. However, previous observational studies yielded inconsistent findings. The present 2-sample Mendelian randomization study provides a suggestive evidence for the causal relationship between statins use and lower risk of COVID-19 hospitalization, which could overcome the limitations of observational study (i.e., confounding bias and reverse causation).

