## Supplements for "Association of lipid-lowering drugs with COVID-19 outcomes: A Mendelian Randomization study"

**SUPPLEMENTARY MATERIALS**

**Contents:**

**Supplementary Table 1.** Information of eQTL and GWAS summary data.

**Supplementary Table 2.** SMR association between expression of gene HMGCR, PCSK9, or NPC1L1 and COVID-19 outcomes.

**Supplementary Table 3.** Information of genetic instrumental variants associated with LDL cholesterol located within 100 kb windows from gene HMGCR, PCSK9 or NPC1L1.

**Supplementary Table 4.** IVW-MR association between LDL cholesterol mediated by gene HMGCR, PCSK9, or NPC1L1and COVID-19 outcomes.

**Supplementary table 5.** SMR association between expression of gene HMGCR, PCSK9, or NPC1L1 and LDL cholesterol level.

**Supplementary table 6.** IVW-MR association between LDL cholesterol mediated by gene HMGCR, PCSK9, or NPC1L1 and coronary heart disease.

**Supplementary Table 7.** Association between eQTL top SNP of HMGCR with expression of other nearby genes (p<0.05).

**Supplementary Table 8.** SMR association between expression of genes adjactent to HMGCR and COVID-19 outcomes.

**Supplementary Table 1**. Information of eQTL and GWAS summary data.

| Characteristic | Resource | Sample size | Population ancestry | Reference | Data download |
|---|---|---|---|---|---|
| **eQTL data** | | | | | |
| eQTL for HMGCR | eQTLGen Consortium | Whole blood: 31684 | Predominantly European | Võsa, U., Claringbould, A.,Franke, L, et al(2018): Unraveling the polygenic architecture of complex traits using blood eQTL meta-analysis. https://www.biorxiv.org/content/10.1101/447367v1 | https://www.eqtlgen.org/cis-eqtls.html |
| eQTL in PCSK9, NPC1L1 | GTExV8 | Whole blood: 755 Adipose Subcutaneous: 663 | Predominantly European | GTEx Consortium, et al (2017): Genetic effects on gene expression across human tissues. https://www.ncbi.nlm.nih.gov/pubmed/29022597/ | https://www.gtexportal.org/home/datasets |
| **GWAS summary data** | | | | | |
| COVID-19 susceptibility | COVID-19 Host Genetics Initiative | Number of cases:14,134 Number of controls: 1,284,876 | European | COVID-19 Host Genetics Initiative (2020).The COVID-19 Host Genetics Initiative, a global initiative to elucidate the role of host genetic factors in susceptibility and severity of the SARS-CoV-2 virus pandemic. https://pubmed.ncbi.nlm.nih.gov/32404885/ | https://www.covid19hg.org/results/r4/ |
| COVID-19 hospitalization | COVID-19 Host Genetics Initiative | Number of cases:6,406 Number of controls: 902,088 | European | COVID-19 Host Genetics Initiative (2020).The COVID-19 Host Genetics Initiative, a global initiative to elucidate the role of host genetic factors in susceptibility and severity of the SARS-CoV-2 virus pandemic. https://pubmed.ncbi.nlm.nih.gov/32404885/ | https://www.covid19hg.org/results/r4/ |
| COVID-19 very severe respiratory disease | COVID-19 Host Genetics Initiative | Number of cases:3,886 Number of controls: 622,265 | European | COVID-19 Host Genetics Initiative (2020).The COVID-19 Host Genetics Initiative, a global initiative to elucidate the role of host genetic factors in susceptibility and severity of the SARS-CoV-2 virus pandemic. https://pubmed.ncbi.nlm.nih.gov/32404885/ | https://www.covid19hg.org/results/r4/ |
| LDL cholesterol | Global Lipids Genetics Consortium | Number of cases:173,082 Number of controls: 2,437,752 | Predominantly European | Cristen J Willer, Ellen M Schmidt, Sebanti Sengupta, et al (2013): Discovery and refinement of loci associated with lipid levels. https://pubmed.ncbi.nlm.nih.gov/24097068/ | http://csg.sph.umich.edu/willer/public/lipids2013/ |
| Coronary heart disease | CARDIoGRAMplusC4D Consortium | Number of cases:60,801 Number of controls: 123,504 | Predominantly European | Majid Nikpay, Anuj Goel, Hong-Hee Won, et al (2015): A comprehensive 1,000 Genomes-based genome-wide association meta-analysis of coronary artery disease. https://pubmed.ncbi.nlm.nih.gov/26343387/ | http://www.cardiogramplusc4d.org/ |

Abbreviations and Acronyms: eQTLs, expression quantitative trait loci; GWAS, genome-wide association study; HMGCR, HMG-CoA reductase; LDL, low-density lipoprotein; NPC1L1, Niemann-Pick C1-Like 1; PCSK9, Proprotein convertase subtilisin/kexin type 9.

**Supplementary Table 2.** SMR association between expression of gene HMGCR, PCSK9, or NPC1L1 and COVID-19 outcomes.

| | | | | | eQTL association | | | | | GWAS association | | | | SMR association | | | HEIDI Test | |
|---|---|---|---|---|---|---|---|---|---|---|---|---|---|---|---|---|---|---|
| **Gene** | **top eQTL SNP** | **Effect allele** | **Other allele** | **Effect allele frequency** | **Tissue** | **beta** | **se** | **p-value** | **F-statistic** | **Outcome** | **beta** | **se** | **p-value** | **beta** | **se** | **p-value** | **p-value** | **Number of SNPs** |
| HMGCR | rs6453133 | G | A | 0.286 | Blood | 0.128 | 0.009 | 1.21E-50 | 223.999 | COVID-19 susceptibility | 0.033 | 0.014 | 1.56E-02 | 0.262 | 0.110 | 1.70 E-02 | 8.59E-03 | 20 |
| HMGCR | rs6453133 | G | A | 0.286 | Blood | 0.128 | 0.009 | 1.21E-50 | 223.999 | COVID-19 hospitalization | 0.041 | 0.017 | 1.71E-02 | 0.323 | 0.137 | 1.85 E-02 | 1.53E-02 | 20 |
| HMGCR | rs6453133 | G | A | 0.286 | Blood | 0.128 | 0.009 | 1.21E-50 | 223.999 | COVID-19 very severe respiratory disease | 0.034 | 0.030 | 2.52E-01 | 0.269 | 0.236 | 2.54 E-01 | 5.38E-01 | 20 |
| PCSK9 | rs472495 | T | G | 0.628 | Blood | 0.187 | 0.025 | 8.79E-14 | 55.620 | COVID-19 susceptibility | -0.032 | 0.013 | 1.41E-02 | -0.172 | 0.074 | 1.97 E-02 | 6.97E-01 | 8 |
| PCSK9 | rs472495 | T | G | 0.628 | Blood | 0.187 | 0.025 | 8.79E-14 | 55.620 | COVID-19 hospitalization | -0.018 | 0.016 | 2.57E-01 | -0.098 | 0.087 | 2.62 E-01 | 7.73E-01 | 8 |
| PCSK9 | rs472495 | T | G | 0.628 | Blood | 0.187 | 0.025 | 8.79E-14 | 55.620 | COVID-19 very severe respiratory disease | -0.031 | 0.029 | 2.90E-01 | -0.164 | 0.156 | 2.95 E-01 | 4.56E-01 | 8 |
| NPC1L1 | rs41279633 | T | G | 0.144 | Adipose Subcutaneous | 0.473 | 0.054 | 1.69E-18 | 77.023 | COVID-19 susceptibility | 0.032 | 0.018 | 8.56E-02 | 0.067 | 0.040 | 9.16 E-02 | 5.85E-02 | 5 |
| NPC1L1 | rs41279633 | T | G | 0.144 | Adipose Subcutaneous | 0.473 | 0.054 | 1.69E-18 | 77.023 | COVID-19 hospitalization | 0.021 | 0.023 | 3.54E-01 | 0.044 | 0.048 | 3.57 E-01 | 6.64E-02 | 5 |
| NPC1L1 | rs41279633 | T | G | 0.144 | Adipose Subcutaneous | 0.473 | 0.054 | 1.69E-18 | 77.023 | COVID-19 very severe respiratory disease | 0.012 | 0.040 | 7.62E-01 | 0.026 | 0.084 | 7.62 E-01 | 4.86E-01 | 5 |

Abbreviations and Acronyms: eQTLs, expression quantitative trait loci; GWAS, genome-wide association study; HEIDI, heterogeneity in dependent instruments; HMGCR, HMG-CoA reductase; LDL, low-density lipoprotein; NPC1L1, Niemann-Pick C1-Like 1; PCSK9, Proprotein convertase subtilisin/kexin type 9; SNP, single-nucleotide polymorphism; SMR, summary-data-based Mendelian Randomization.

**Supplementary Table 3.** Information of genetic instrumental variants associated with LDL cholesterol located within 100 kb windows from gene HMGCR, PCSK9 or NPC1L1.

| SNP | Effect allele | Other allele | Effect allele frequency | beta | se | p value | F-statistic |
|---|---|---|---|---|---|---|---|
| **HMGCR** | | | | | | | |
| rs12916 | C | T | 0.4314 | 0.0733 | 0.0038 | 7.79E-78 | 372.0838 |
| rs3804231 | A | G | 0.1319 | 0.0642 | 0.0053 | 1.88E-29 | 146.7298 |
| rs10515198 | A | G | 0.1029 | 0.0599 | 0.0061 | 5.99E-22 | 96.42596 |
| rs10066707 | A | G | 0.4169 | 0.0497 | 0.0054 | 2.97E-19 | 84.70816 |
| rs12659791 | C | T | 0.1557 | 0.0433 | 0.005 | 1.42E-18 | 74.9956 |
| rs72633962 | C | T | 0.1412 | 0.06 | 0.0072 | 3.33E-15 | 69.44444 |
| rs3857388 | C | T | 0.1280 | 0.0421 | 0.0059 | 2.20E-11 | 50.91669 |
| **PCSK9** | | | | | | | |
| rs11591147 | T | G | 0.0172 | -0.497 | 0.018 | 8.57E-143 | 762.3735 |
| rs11206510 | C | T | 0.1544 | -0.0831 | 0.005 | 2.38E-53 | 276.2244 |
| rs2479409 | A | G | 0.6675 | -0.0642 | 0.0041 | 2.51E-50 | 245.1898 |
| rs585131 | T | C | 0.8153 | 0.0637 | 0.005 | 2.70E-35 | 162.3076 |
| rs11206514 | A | C | 0.6108 | 0.0507 | 0.0041 | 9.95E-33 | 152.9143 |
| rs572512 | T | C | 0.3456 | 0.0478 | 0.0047 | 5.31E-26 | 103.4332 |
| rs2479394 | A | G | 0.7150 | -0.0386 | 0.0041 | 1.58E-19 | 88.63534 |
| rs12067569 | A | G | 0.0343 | 0.0885 | 0.01 | 1.97E-17 | 78.3225 |
| rs10493176 | G | T | 0.1148 | -0.0776 | 0.0102 | 2.54E-14 | 57.87928 |
| rs4927193 | C | T | 0.1306 | -0.0352 | 0.0056 | 4.27E-11 | 39.5102 |
| rs11583974 | A | G | 0.0303 | 0.0646 | 0.0117 | 3.95E-09 | 30.4855 |
| rs2495495 | C | T | 0.8654 | -0.0342 | 0.0059 | 3.52E-08 | 33.60069 |
| **NPC1L1** | | | | | | | |
| rs2073547 | G | A | 0.1939 | 0.0485 | 0.0049 | 1.92E-21 | 97.9696 |
| rs217386 | A | G | 0.4077 | -0.0363 | 0.0038 | 1.20E-19 | 91.25277 |
| rs7791240 | C | T | 0.0910 | 0.0425 | 0.0065 | 1.84E-10 | 42.75148 |

Abbreviations and Acronyms: HMGCR, HMG-CoA reductase; LDL, low-density lipoprotein; NPC1L1, Niemann-Pick C1-Like 1; PCSK9, Proprotein convertase subtilisin/kexin type 9; SNP, single-nucleotide polymorphism.

**Supplementary Table 4.** IVW-MR association between LDL cholesterol mediated by gene HMGCR, PCSK9, or NPC1L1and COVID-19 outcomes.

| Exposure | Outcome | beta | se | p value | p value for Cochran Q test | p value for MR-Egger intercept | p value for MR-PRESSO Global test |
|---|---|---|---|---|---|---|---|
| HMGCR | COVID-19 susceptibility | 0.082902782 | 0.112458917 | 0.461 | 0.654 | 0.942 | 0.716 |
| HMGCR | COVID-19 hospitalization | 0.277597811 | 0.141059053 | 0.049 | 0.891 | 0.678 | 0.908 |
| HMGCR | COVID-19 very severe respiratory disease | 0.211069627 | 0.24369484 | 0.386 | 0.665 | 0.722 | 0.699 |
| PCSK9 | COVID-19 susceptibility | -0.0114243 | 0.071333551 | 0.873 | 0.917 | 0.284 | 0.843 |
| PCSK9 | COVID-19 hospitalization | 0.0265409 | 0.08529678 | 0.756 | 0.936 | 0.116 | 0.77 |
| PCSK9 | COVID-19 very severe respiratory disease | -0.01812229 | 0.172247404 | 0.916 | 0.714 | 0.498 | 0.727 |
| NPC1L1 | COVID-19 susceptibility | 0.702007394 | 0.224546332 | 0.002 | 0.752 | 0.814 | N/A |
| NPC1L1 | COVID-19 hospitalization | 0.517697157 | 0.279906967 | 0.064 | 0.830 | 0.700 | N/A |
| NPC1L1 | COVID-19 very severe respiratory disease | 0.704499377 | 0.477430503 | 0.140 | 0.372 | 0.396 | N/A |

Abbreviations and Acronyms: HMGCR, HMG-CoA reductase; IVW-MR, inverse-variance weighted Mendelian Randomization; LDL, low-density lipoprotein; MR-PRESSO, Mendelian Randomization Pleiotropy RESidual Sum and Outlier; NPC1L1, Niemann-Pick C1-Like 1; PCSK9, Proprotein convertase subtilisin/kexin type 9; SNP, single-nucleotide polymorphism.

**Supplementary table 5.** SMR association between expression of gene HMGCR, PCSK9, or NPC1L1 and LDL cholesterol level.

| Exposure | OR | 95%CI | p value | Tissue |
|---|---|---|---|---|
| HMGCR | 1.48 | 1.36-1.6 | 1.18E-21 | Blood |
| PCSK9 | 1.45 | 1.28-1.63 | 2.77E-09 | Blood |
| NPC1L1 | 1.11 | 1.07-1.16 | 1.30E-08 | Adipose Subcutaneous |

Abbreviations and Acronyms: CI, confidence interval; HMGCR, HMG-CoA reductase; LDL, low-density lipoproteinNPC1L1, Niemann-Pick C1-Like 1; OR, odds ratio; PCSK9, Proprotein convertase subtilisin/kexin type 9; SMR, summary-data-based Mendelian Randomization.

**Supplementary table 6.** IVW-MR association between LDL cholesterol mediated by gene HMGCR, PCSK9, or NPC1L1 and coronary heart disease.

| Exposure | OR | 95%CI | p value |
|---|---|---|---|
| HMGCR | 1.44 | 1.24-1.68 | 2.18E-06 |
| PCSK9 | 1.67 | 1.44-1.92 | 3.54E-12 |
| NPC1L1 | 1.66 | 1.2-2.28 | 2.07E-03 |

Abbreviations and Acronyms: CI, confidence interval; HMGCR, HMG-CoA reductase; IVW-MR, inverse-variance weighted Mendelian Randomization; LDL, low-density lipoproteinNPC1L1, Niemann-Pick C1-Like 1; OR, odds ratio; PCSK9, Proprotein convertase subtilisin/kexin type 9.

**Supplementary Table 7.** Association between eQTL top SNP of HMGCR (rs6453133) with expression of other nearby genes (p<0.05).

| **Gene** | **eQTL SNP** | **Effect allele** | **Other allele** | **Effect allele frequency** | **eQTL association** | | |
|---|---|---|---|---|---|---|---|
| | | | | | **beta** | **se** | **p-value** |
| POC5 | rs6453133 | G | A | 0.31301 | 0.16121 | 0.00855 | 2.43E-79 |
| HMGCR | rs6453133 | G | A | 0.31301 | 0.12776 | 0.00854 | 1.21E-50 |
| ANKDD1B | rs6453133 | G | A | 0.31301 | -0.0354 | 0.01029 | 0.00059 |
| COL4A3BP | rs6453133 | G | A | 0.31301 | -0.0205 | 0.00938 | 0.02854 |
| POLK | rs6453133 | G | A | 0.31301 | 0.01854 | 0.00857 | 0.03046 |
| ANKRD31 | rs6453133 | G | A | 0.31301 | -0.0228 | 0.01161 | 0.04905 |

Abbreviations and Acronyms: eQTLs, expression quantitative trait loci; HMGCR, HMG-CoA reductase.

**Supplementary Table 8.** SMR association between expression of genes adjactent to HMGCR and COVID-19 outcomes.

| Outcome | Gene | top eQTL SNP | Effect allele | Other allele | Effect allele frequency | eQTL association beta | eQTL association se | eQTL association p-value | eQTL association F-statistic | GWAS association beta | GWAS association se | GWAS association p-value | SMR association beta | SMR association se | SMR association p-value | HEIDI Test p-value | HEIDI Test Number of SNPs |
|---|---|---|---|---|---|---|---|---|---|---|---|---|---|---|---|---|---|
| COVID-19 susceptibility | HMGCR | rs6453133 | G | A | 0.286 | 0.128 | 0.009 | 1.21E-50 | 223.999 | 0.033 | 0.014 | 1.56E-02 | **0.262** | **0.110** | **1.70E-02** | 8.59E-03 | 20 |
| COVID-19 susceptibility | COL4A3BP | rs10515198 | A | G | 0.101 | -0.119 | 0.014 | 8.07E-17 | 69.392 | -0.022 | 0.022 | 3.09E-01 | 0.186 | 0.184 | 3.12E-01 | 1.76E-01 | 20 |
| COVID-19 susceptibility | ANKDD1B | rs6895057 | A | G | 0.259 | -0.074 | 0.011 | 8.61E-12 | 46.622 | -0.007 | 0.014 | 6.07E-01 | 0.099 | 0.194 | 6.08E-01 | 1.80E-01 | 8 |
| COVID-19 susceptibility | POC5 | rs113216064 | T | C | 0.184 | 0.565 | 0.009 | 0.00E+00 | 3743.448 | 0.005 | 0.016 | 7.51E-01 | 0.009 | 0.028 | 7.51E-01 | 4.50E-01 | 20 |
| COVID-19 hospitalization | HMGCR | rs6453133 | G | A | 0.286 | 0.128 | 0.009 | 1.21E-50 | 223.999 | 0.041 | 0.017 | 1.71E-02 | **0.323** | **0.137** | **1.85E-02** | 1.53E-02 | 20 |
| COVID-19 hospitalization | COL4A3BP | rs10515198 | A | G | 0.101 | -0.119 | 0.014 | 8.07E-17 | 69.392 | 0.009 | 0.027 | 7.49E-01 | -0.072 | 0.225 | 7.49E-01 | 4.28E-01 | 20 |
| COVID-19 hospitalization | ANKDD1B | rs6895057 | A | G | 0.259 | -0.074 | 0.011 | 8.61E-12 | 46.622 | -0.001 | 0.018 | 9.64E-01 | 0.011 | 0.238 | 9.64E-01 | 2.46E-01 | 8 |
| COVID-19 hospitalization | POC5 | rs113216064 | T | C | 0.184 | 0.565 | 0.009 | 0.00E+00 | 3743.448 | 0.029 | 0.020 | 1.44E-01 | 0.051 | 0.035 | 1.44E-01 | 2.22E-02 | 20 |
| COVID-19 very severe respiratory disease | HMGCR | rs6453133 | G | A | 0.286 | 0.128 | 0.009 | 1.21E-50 | 223.999 | 0.034 | 0.030 | 2.52E-01 | 0.269 | 0.236 | 2.54E-01 | 5.38E-01 | 20 |
| COVID-19 very severe respiratory disease | COL4A3BP | rs10515198 | A | G | 0.101 | -0.119 | 0.014 | 8.07E-17 | 69.392 | -0.049 | 0.046 | 2.96E-01 | 0.408 | 0.393 | 3.00E-01 | 2.23E-01 | 20 |
| COVID-19 very severe respiratory disease | ANKDD1B | rs6895057 | A | G | 0.259 | -0.074 | 0.011 | 8.61E-12 | 46.622 | -0.005 | 0.031 | 8.85E-01 | 0.061 | 0.420 | 8.85E-01 | 6.40E-01 | 8 |
| COVID-19 very severe respiratory disease | POC5 | rs113216064 | T | C | 0.184 | 0.565 | 0.009 | 0.00E+00 | 3743.448 | -0.006 | 0.035 | 8.57E-01 | -0.011 | 0.062 | 8.57E-01 | 7.71E-01 | 20 |

Abbreviations and Acronyms: eQTLs, expression quantitative trait loci; GWAS, genome-wide association study; HEIDI, heterogeneity in dependent instruments; HMGCR, HMG-CoA reductase; SNP, single-nucleotide polymorphism; SMR, summary-data-based Mendelian Randomization.

*No eQTLs for POLK or ANKRD31 are available at a genome-wide significance level (p < 5.0 × 10-8).
